# Suppression of Endogenous Alpha Power Predicts Clinical Response to 10 Hz tACS in Major Depressive Disorder: A Double-Blind Randomized Controlled Trial

**DOI:** 10.64898/2026.03.17.26348625

**Authors:** Tobias Schwippel, Francesca Pupillo, Hadden LaGarde, Athena Stein, Mengsen Zhang, David Rubinow, Flavio Frohlich

## Abstract

**Background:** Transcranial alternating current stimulation (tACS) is a promising non-pharmacological intervention for major depressive disorder (MDD), but its effects on endogenous alpha oscillatory dynamics and their relationship to clinical improvement remain unclear.

**Methods:** In this double-blind, sham-controlled randomized clinical trial, 20 adults with MDD received five consecutive days of prefrontal 10 Hz tACS or sham. Resting 128-channel EEG was acquired before stimulation on Day 1 (D1), Day 5 (D5), and two-week follow-up. Changes in alpha power spectral density were quantified at the stimulation frequency (10 Hz) and at each participant’s individual alpha frequency (IAF), using prefrontal regions of interest and whole-head topographical analyses. Depression severity was assessed using the Hamilton Depression Rating Scale (HDRS-17).

**Results:** Between-group comparisons revealed no significant differences in prefrontal alpha power changes at either 10 Hz or IAF during the intervention week or at follow-up, although right prefrontal 10 Hz power showed a trend-level reduction with tACS. In contrast, within the tACS group, greater reductions in prefrontal IAF power were associated with greater HDRS-17 improvement from D1 to follow-up, and early IAF power suppression during the intervention week predicted later symptom improvement. Whole-head analyses identified a posterior cluster of reduced 10 Hz power at follow-up in the tACS group relative to sham, whereas clinically relevant correlations were specific to IAF power and distributed across frontal-central and parietal electrodes. Depression scores improved over time in both groups, with greater reductions in HDRS-17 scores observed in the tACS group.

**Conclusions:** Findings suggest that five days of 10 Hz tACS engages depression-relevant alpha mechanisms, with symptom improvement linked specifically to modulation of alpha power at IAF. Results support personalization of tACS in future trials.

## Introduction

Major depressive disorder (MDD) is a highly prevalent and disabling condition that contributes substantially to global years lived with disability [1]. Despite available pharmacotherapies and psychotherapies, real-world effectiveness is constrained by delayed onset, incomplete response, adverse effects, and limited access. The resulting treatment gap is large: cross-national survey data indicate that only a minority of individuals meeting criteria for MDD receive minimally adequate care [2]. These challenges motivate the development of safe, scalable, and mechanistically grounded interventions that can be tailored to individual neurobiology.

MDD is increasingly conceptualized as a disorder of large-scale brain network organization [3]. In the resting brain, alpha-band activity (∼8–12 Hz) is the dominant EEG rhythm and is thought to implement functional inhibition, regulating information flow through rhythmic gating of cortical excitability [4]. Consistent with this framework, alpha power has been linked to reduced local cortical excitability and is frequently anticorrelated with fMRI BOLD activity [5]. At the scalp level, resting-state alpha oscillations are most prominent over occipital and parietal regions, where they have been associated with internally oriented processing [6], and extend to frontal areas involved in cognitive control.

Alterations in alpha oscillations have been consistently implicated in MDD. Early work primarily emphasized increased prefrontal alpha power and frontal alpha power asymmetry [7], while more recent evidence indicates that these abnormalities extend to posterior cortical areas [8]. Elevated alpha power in depression may index maladaptive inhibitory dynamics that constrain flexible coordination across distributed networks, potentially stabilizing maladaptive brain states. Clinically, such dynamics could support excessive internally directed processing and reduced cognitive flexibility.

Transcranial alternating current stimulation (tACS) perturbs ongoing oscillations by applying oscillatory electric fields delivered at controlled frequencies [9]. In driven systems, entrainment emerges within a frequency–intensity window (Arnold tongue), such that external stimulation preferentially engages endogenous rhythms when closely matched in frequency [10]. At the neuronal level, weak electric fields bias spike timing and modulate ongoing oscillations [11], effects that scale to large-scale networks as resonance phenomena [12]. These dynamics likely reflect a combination of transient entrainment and longer-lasting spike-timing–dependent plasticity [13].

A previous randomized controlled trial reported that five days of 10 Hz tACS targeting the dorsolateral prefrontal cortex reduced depressive symptoms and were accompanied by reductions in frontal alpha power [14]. These findings provided initial evidence that externally applied oscillatory stimulation can engage alpha-band dynamics relevant to depression. However, the specific mechanisms underlying these effects remain unclear. In particular, it is unresolved whether clinical improvement reflects modulation of power at the applied stimulation frequency (10 Hz) or changes in endogenous alpha oscillations indexed by power at the individual alpha frequency (IAF). In addition, clinical improvements in that study emerged most prominently at follow-up, raising the possibility that repeated stimulation induces delayed plasticity-related changes in alpha oscillatory dynamics rather than purely acute entrainment effects. Finally, the spatial extent of such modulation remains uncertain, ranging from local effects near stimulation sites to distributed changes consistent with large-scale network engagement.

Against this background, we conducted a double-blind, sham-controlled randomized trial to test whether five days of bifrontal-central 10 Hz tACS modulates endogenous alpha oscillations and whether such modulation relates to clinical improvement. Using repeated 128-channel EEG, we focused on the two-week follow-up and examined the frequency specificity, temporal dynamics, and cortical distribution of stimulation effects. Based on prior evidence linking tACS to reductions in alpha power, we hypothesized that clinical improvement at the two-week follow-up would be associated with reductions in alpha power. We further examined whether these changes occurred at the stimulation frequency (10 Hz) or at the IAF.

## Methods

### Study Design and Experimental Procedures

This randomized, sham-controlled, double-blind, parallel-design clinical trial investigated the effects of five-day 10 Hz tACS on depression symptoms in MDD (NCT03994081). The study was conducted at the Carolina Center for Neurostimulation at Chapel Hill, North Carolina between June 2021 and June 2023. All participants provided written informed consent prior to enrollment and the study was approved by the University of North Carolina-Chapel Hill Office of Human Research Ethics (IRB # 20-1822). After remote prescreening and eligibility confirmation via remote interview, participants received five consecutive daily sessions of either 10 Hz tACS or sham stimulation. Clinical assessments and high-density EEG (HD-EEG) recordings were obtained on Day 1 (D1), Day 5 (D5), and at a two-week follow-up (FU). The primary outcomes were (1) changes in alpha power spectral density (PSD) (D5-D1, FU-D1) and (2) their association with corresponding changes in depression severity according to the Hamilton Depression Rating Scale (HDRS-17).

### Participants

Twenty participants were enrolled (see CONSORT flow diagram, Supplementary Figure S1). Inclusion criteria were: age 18-70 years; DSM-IV diagnosed unipolar, non-psychotic MDD and HDRS-17 scores >8. Low suicide risk was confirmed via the C-SSRS. Exclusion criteria were: diagnosis of moderate or severe alcohol/substance use disorder excluding tobacco (within the last 12 months); bipolar or psychotic disorder (lifetime); eating disorder (within the last six months); obsessive-compulsive disorder (lifetime); post-traumatic stress disorder (within the last six months); prior brain surgery, brain implants; current pregnancy; change of antidepressant medication within the last four weeks; newly initiated antidepressant within the last four weeks; non-English speakers.

### Clinical Assessments

Depression severity was assessed using the clinician-rated HDRS-17 [15] and the self-reported Beck-Depression-Inventory (BDI-II) [16]. Additional questionnaires measured anxiety (State-Trait-Anxiety-Inventory, STAI) [17], anhedonia (Snaith-Hamilton-Pleasure-Scale, SHAPS) [18], rumination (Rumination Response Scale, RSS) [19], depression and anxiety (Inventory of Depression and Anxiety Symptoms, IDAS) [20], and quality of life (Quality of Life Enjoyment and Satisfaction Questionnaire-Short Form, Q-LES-Q-SF) [21] at D1, D5, and FU. Due to a data storage failure, data from the additional self-report questionnaires (STAI, SHAPS, IDAS, Q-LES-Q-SF) were unavailable for participants 1–5, while BDI-II data remained intact. Consequently, analyses involving these measures were conducted in a reduced sample (D1: sham n = 10, tACS n = 10; D5: sham n = 8, tACS n = 7; FU: sham n = 8, tACS n = 7).

### Transcranial Alternating Current Stimulation

tACS was delivered via the XCSITE device (Pulvinar Neuro, Durham, NC) at 10 Hz, 2 mA zero-to-peak. Two 5 × 5 cm carbon-silicone electrodes were positioned over the left (F3) and right (F4) prefrontal cortex, and a 5 × 7 cm electrode was placed at the vertex (Cz). The current between the prefrontal electrodes was split, delivering 1 mA in-phase stimulation to each prefrontal site, while the vertex received 2 mA anti-phase stimulation. Each session lasted 40 minutes, including 20 s ramp-up and ramp-down. Active sham stimulation consisted of 20 s ramp-up, 40 s of 10 Hz tACS, and 20 s ramp-down. During stimulation, participants watched a relaxing video (ReefScapes, Undersea Productions, Queensland, Australia) to standardize visual input for effective blinding, as well as brain state. Randomization was computer-generated by an independent researcher (M.Z.) with no participant contact. Participants and outcome assessors were blinded to stimulation conditions.

### EEG Recording and Preprocessing

Resting-state, eyes-open high-density EEG (HD-EEG) was recorded daily before and after stimulation at 1000 Hz using a 128-channel system (NetAmps 410, Electrical Geodesics, Inc, Eugene, Oregon). For the present study, analyses focused on recordings obtained before stimulation at D1 and D5 and at FU. Analyses of within-week changes during the intervention period are reported elsewhere (Stein & Schwippel et al., submitted). Data were imported into MATLAB via EEGLAB [22], bandpass filtered between 1–40 Hz, and downsampled to 200 Hz. Noisy channels were manually identified and interpolated, and data were re-referenced to the common average. Independent component analysis (ICA) was performed, and components were classified using the ICLabel algorithm. Components were rejected when class confidence exceeded 0.5 for muscle, eye, heart, or channel noise and 0.8 for line noise; components classified as brain or “other” were retained [23]. PSD was computed using Welch’s method, and the FOOOF toolbox was used to decompose the PSD into periodic and aperiodic components (2–40 Hz; maximum of three peaks; fixed aperiodic mode) [24]. The aperiodic component was subtracted to obtain the flattened PSD, which was used to extract PSD values at the stimulation frequency (10 Hz) and at each participant’s daily peak individual alpha frequency (IAF). Regions of interest (ROIs) were defined by six electrodes surrounding and including F3 and F4, covering the cortical regions targeted by tACS.

### Statistical Analyses

Analyses were conducted in R 4.2 [25] and MATLAB R2025a (The MathWorks, Inc., Natick, Massachusetts, USA). Statistical significance was set at α = 0.05 (two-tailed). EEG analyses focused on alpha PSD at 10 Hz and IAF. Given the sample size, nonparametric methods were used for EEG group comparisons and correlations. A detailed statistical analysis plan is provided in the Supplement.

#### Primary Analyses

Prefrontal alpha PSD change scores (D5–D1 and FU–D1) were computed separately for the tACS and sham groups and analyzed using Wilcoxon signed-rank tests within each group. Associations between PSD changes and HDRS-17 change scores were assessed using Spearman correlations. These ROI-based analyses correspond to the pre-registered primary outcomes (https://ClinicalTrials.gov NCT03994081).

#### Secondary Analyses

Whole-head PSD changes and their associations with clinical improvement were evaluated using nonparametric cluster-based permutation testing with family-wise error correction, and channel-wise correlations controlled using false discovery rate (FDR) procedures (see Supplementary Statistical Methods for full specification). Clinical outcomes (HDRS-17, BDI-II) were analyzed using two-way repeated-measures ANOVA with Greenhouse–Geisser correction where appropriate. Outcomes with missing observations were analyzed using linear mixed-effects models with condition, time (modeled as a categorical factor), and their interaction as fixed effects, and participant-level random intercepts [26]. Models were estimated using maximum likelihood, and p-values were obtained using Satterthwaite’s approximation. Where appropriate, post hoc comparisons were conducted using estimated marginal means with Holm correction for multiple testing [27]. Treatment response (≥50% HDRS-17 reduction) and remission (HDRS-17 < 8 at FU) were compared between groups using two-sided Fisher’s exact tests. Anticipated stimulation-related sensations were rated after each session on a 4-point scale (0–3). Between-group differences in occurrence and mean severity were analyzed using Fisher’s exact tests and independent-sample t-tests.

## Results

### Participants

Between August 2021 and May 2023, 20 participants were randomized to receive either 10 Hz tACS or sham stimulation. All participants completed the 5-day intervention, and one tACS participant was lost to follow-up. Baseline demographics, medication status, treatment resistance, and depression severity did not differ significantly between groups (Table S1). The sham group had significantly longer current depressive episodes (M = 10.78 vs. 3.15 months, t(18) = 2.81, p = 0.012) and a higher proportion of individuals receiving psychotherapy (88% vs. 20%, χ²(1, N = 19) = 6.49, p = .011).

### Prefrontal ROI Analysis: Alpha PSD Change

No significant group differences were observed in alpha PSD change at 10 Hz or IAF for either prefrontal ROI within the intervention week (D5–D1 all p > 0.121). Likewise, alpha PSD changes between D1 and FU did not differ significantly between conditions. A trend-level effect was observed for 10 Hz PSD in the right prefrontal ROI, with a numerically greater decrease in the tACS group compared with sham, consistent with the hypothesized direction (Z = 68, p = 0.066). Descriptively, prefrontal alpha PSD increased across the intervention week in the sham group, whereas it remained stable in the tACS group. Conversely, from D1 to FU, alpha PSD numerically decreased in the tACS group but not in sham (Figure 1).

**Figure 1.**
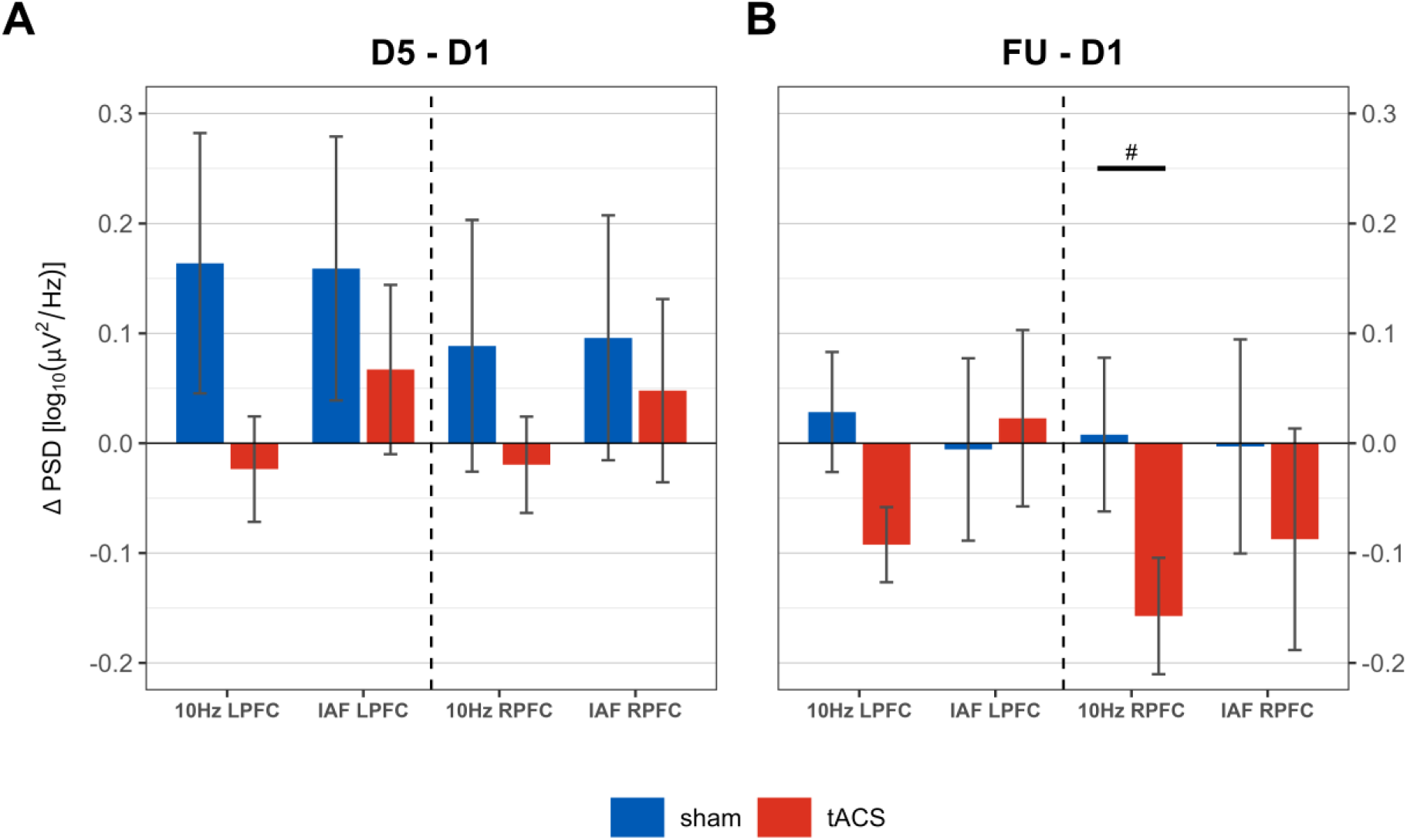
Target Engagement: ROI Analysis of Alpha PSD Modulation. Difference scores for left (LPFC) and right (RPFC) prefrontal electrode seeds have been calculated for IAF and 10 Hz PSD respectively. **A**: Alpha PSD change within the intervention week (D5-D1); **B**: Alpha PSD Change from Day 1 to 2-week follow-up (FU-D1). Error bars represent standard error of the mean. #: p = 0.066.

### Prefrontal ROI Analysis: Alpha PSD and HDRS-17 Change

Changes in prefrontal alpha PSD (at 10 Hz and IAF) during the intervention week (D5–D1) were not significantly correlated with concurrent HDRS-17 changes over the same interval in both conditions. In contrast, the tACS group demonstrated significant and directionally consistent correlations from D1 to FU, with greater bilateral prefrontal IAF PSD decreases associated with greater symptom reduction, consistent with the a priori hypothesis (left: ρ = 0.81, p = 0.008; right: ρ = 0.67, p = 0.0498; Figure 2 C/D). The sham group did not exhibit significant correlations (left: ρ = 0.24, p = 0.5; right ρ = 0.11, p = 0.76).

**Figure 2.**
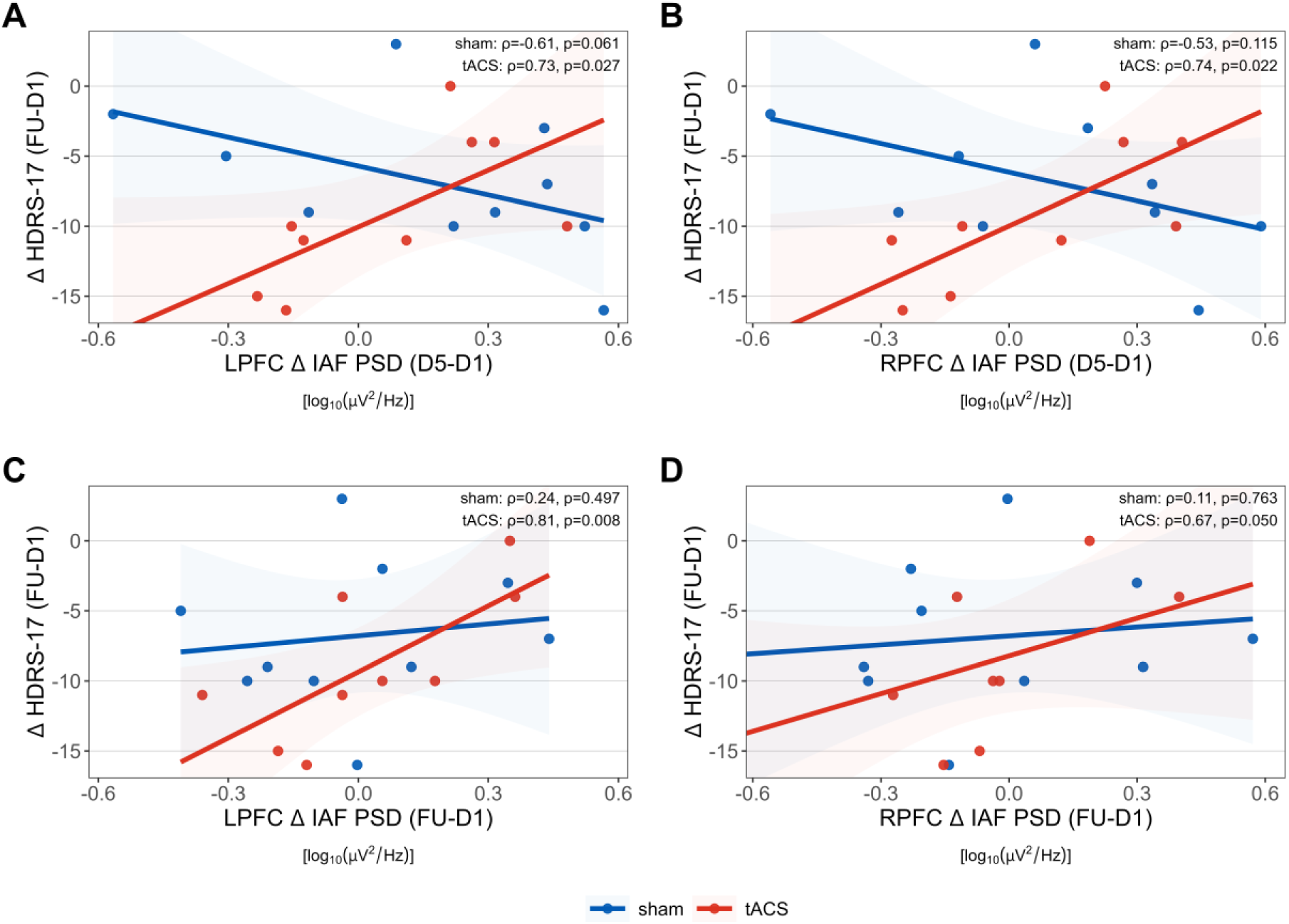
Target Validation: Early and Sustained Prefrontal Alpha Modulation Predicts Depressive Symptom Improvement at 2-Week Follow-Up. **A–D:** Spearman correlations between changes in prefrontal alpha power at the individual alpha frequency (IAF PSD) and depressive symptom change (HDRS-17), shown separately for sham and tACS groups. Panels **A (LPFC)** and **B (RPFC):** IAF PSD change during the intervention week (D5-D1) in relation to HDRS-17 change (FU-D1). Panels **C (LPFC)** and **D (RPFC):** IAF PSD change from Day 1 to follow-up (FU-D1) in relation to HDRS-17 change (FU-D1). Lines represent linear regression fits with shaded 95% confidence intervals, stratified by stimulation condition. Spearman correlation coefficients (ρ) and two-tailed p values are shown within each panel. <u>Abbreviations</u>: IAF: individual alpha frequency; PSD: power spectral density; HDRS-17: Hamilton Depression Rating Scale; tACS: transcranial alternating current stimulation; LPFC/RPFC, left/right prefrontal cortex; D1: Day 1; D5: Day 5; FU: 2-week follow-up.

Additionally, greater bifrontal IAF PSD reductions during the intervention week (D5-D1) were associated with greater FU-D1 HDRS-17 improvement in the tACS group (left: ρ = 0.73, p = 0.027; right: ρ = 0.74, p = 0.022; Figure 2 A/B), whereas correlations in the sham group were directionally opposite and non-significant. Importantly, analogous analyses using 10 Hz PSD did not yield significant correlations in either group, indicating that the observed association between alpha power change and symptom improvement was specific to IAF PSD.

### Topographical Analysis: Alpha PSD Change

Cluster permutation testing of whole-brain alpha PSD changes (at 10 Hz and IAF) during the intervention week revealed no significant differences between the tACS and sham groups. However, examination of 10 Hz PSD changes from D1 to FU identified one significant negative cluster (clustermass = −57.796, p = .032) encompassing 23 occipital and central electrodes indicating a marked reduction of 10 Hz PSD from D1 to FU in the tACS group compared with sham (Figure 3, S2 for IAF PSD results).

**Figure 3.**
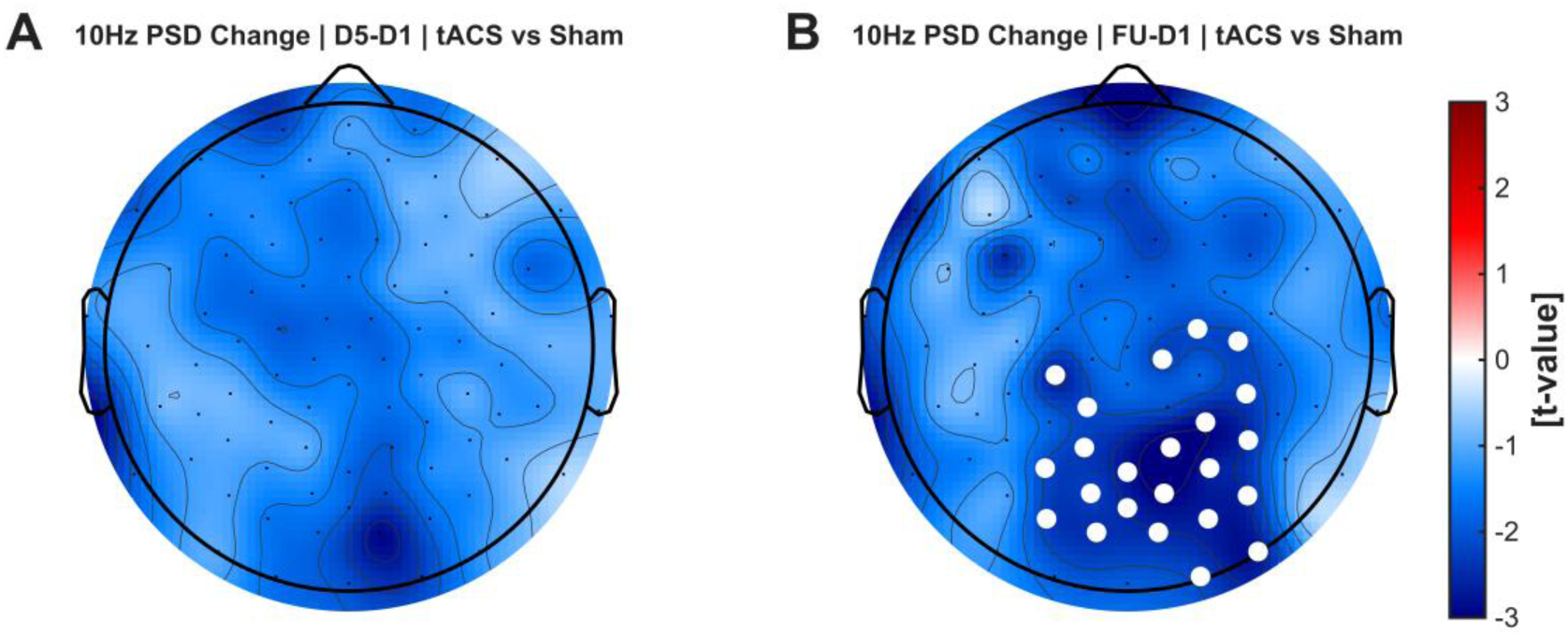
Target Engagement: Topographical Analysis of 10 Hz PSD Modulation. Scalp distributions of t-values derived from nonparametric cluster-based permutation tests comparing PSD change between tACS and sham (double contrast: ΔtACS − Δsham). **A:** Change of 10 Hz PSD from D1 to D5 (intervention week). **B:** Change of 10 Hz PSD from D1 to two-week FU. Colored maps display channel-wise t-statistics, scaled symmetrically around zero. Electrodes marked with white circles denote channels belonging to clusters surviving cluster-level FWER correction (p < .05). Negative t-values indicate greater reductions in 10 Hz PSD in the tACS group relative to sham. Black dots represent EEG electrode locations. <u>Abbreviations</u>: PSD: power spectral density; tACS: transcranial alternating current stimulation; D1: Day 1; D5: Day 5; FU: 2-week follow-up.

### Topographical Analysis: Alpha PSD and HDRS-17 Change

During the intervention week, separate whole-brain channel-wise correlations for the tACS and sham groups revealed no significant associations between alpha PSD change (at 10 Hz and IAF) and HDRS-17 change (Figure 4A, S3 for 10 Hz PSD results). In contrast, within the tACS group, reductions in IAF PSD from D1 to FU were correlated with HDRS-17 improvement from D1 to FU. After FDR correction, significant correlations (p_FDR_ < .05) were observed at 24 electrodes, predominantly over frontal-central and left parietal regions. Spearman coefficients ranged from 0.80 to 0.93, reflecting a strong association between greater alpha PSD reductions and larger depression symptom improvement (Figure 4C).

**Figure 4.**
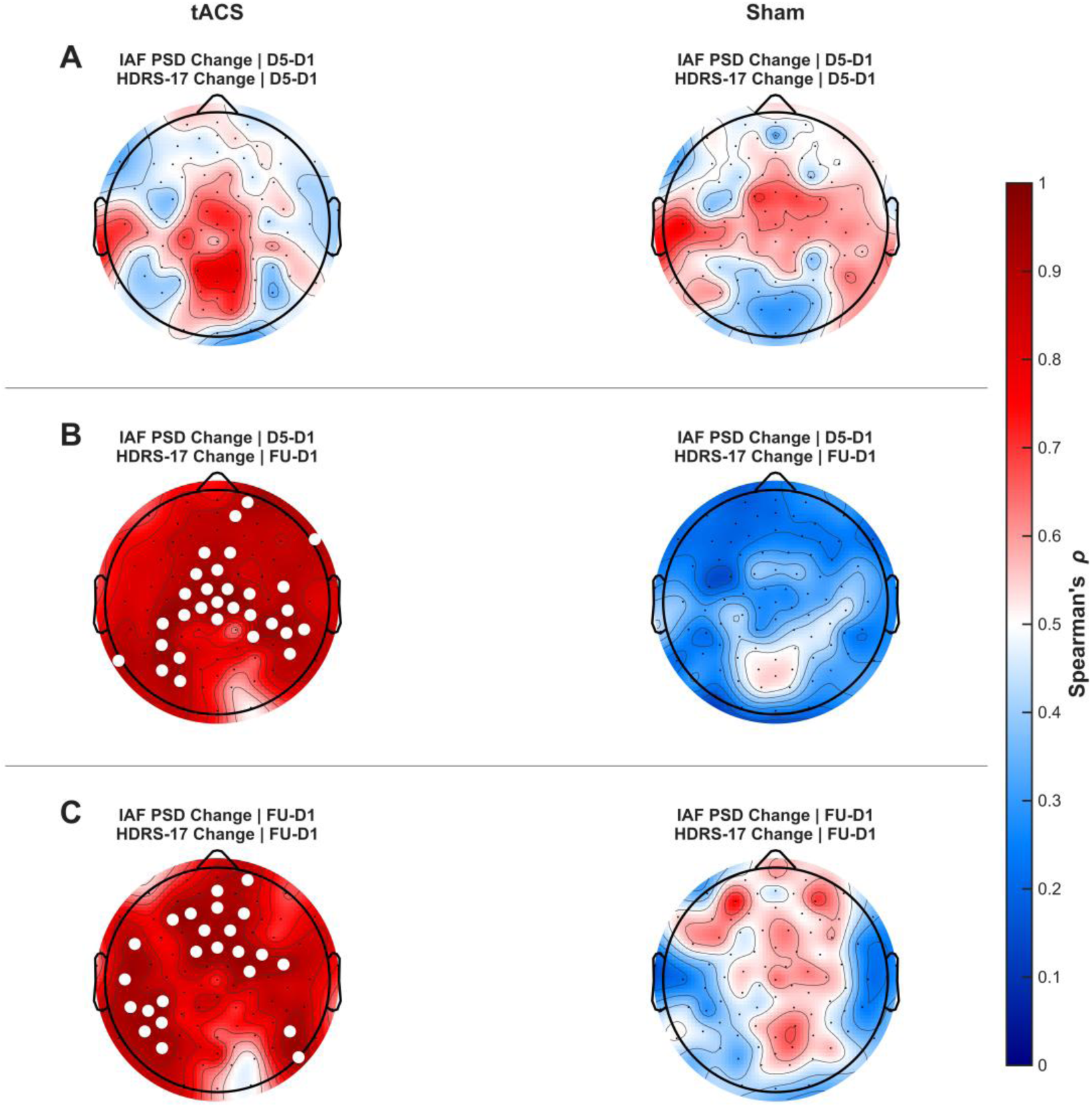
Target Validation: Early and Sustained IAF PSD Reduction Correlates with Symptom Improvement. Topographical maps depict Spearman correlation coefficients between changes in IAF PSD and changes in HDRS-17 across EEG channels, shown separately for tACS and sham groups. Colors indicate correlation coefficients, with positive values reflecting greater symptom reduction associated with larger PSD decreases. White circles indicate FDR-corrected significant channels (q < .05). Black dots indicate EEG electrode locations. **A:** Correlation between IAF PSD change during the intervention week (D5–D1) and concurrent HDRS-17 change (D5–D1). **B:** Correlation between IAF PSD change during the intervention week (D5–D1) and subsequent clinical improvement at 2-week follow-up (FU–D1). **C:** Correlation between sustained PSD change from D1 to FU and HDRS-17 change from D1 to FU. <u>Abbreviations</u>: PSD: power spectral density; tACS: transcranial alternating current stimulation; D1: Day 1; D5: Day 5; FU: 2-week follow-up.

We next examined whether IAF PSD changes during the intervention week were associated with clinical improvement at FU by correlating D5-D1 PSD changes with FU-D1 HDRS-17 changes. In the tACS group, significant correlations emerged across 30 central and bilateral parietal electrodes (p_FDR_ < .05; Spearman ρ = 0.79 to 0.93), with greater IAF PSD reductions associated with greater symptom improvement at 2-week FU (Figure 4B). No significant effects were observed in the sham group, where correlation coefficients were nonsignificant and in the opposite direction.

### Clinical Outcome

RM-ANOVAs revealed a significant main effect of time on both HDRS-17 (F(2,34) = 29.16, p < 0.001, ηp² = .63) and BDI-II (F(2,34) = 23.7, p < 0.001, ηp² = .58), indicating substantial overall symptom improvement with large effect sizes (Figure 5 A/B). No significant time × condition interactions were observed, indicating similar clinical trajectories across groups. At follow-up, reductions in depression scores were numerically larger in the tACS group for HDRS-17 (-9.0 in tACS vs. -6.8 in sham) and BDI-II (-10.3 vs. -8.7). Response rates (55.6% in tACS vs. 60% in sham) and remission rates (33.3% vs. 40%) did not differ between groups (Fisher’s exact test, both p = 1.00).

**Figure 5.**
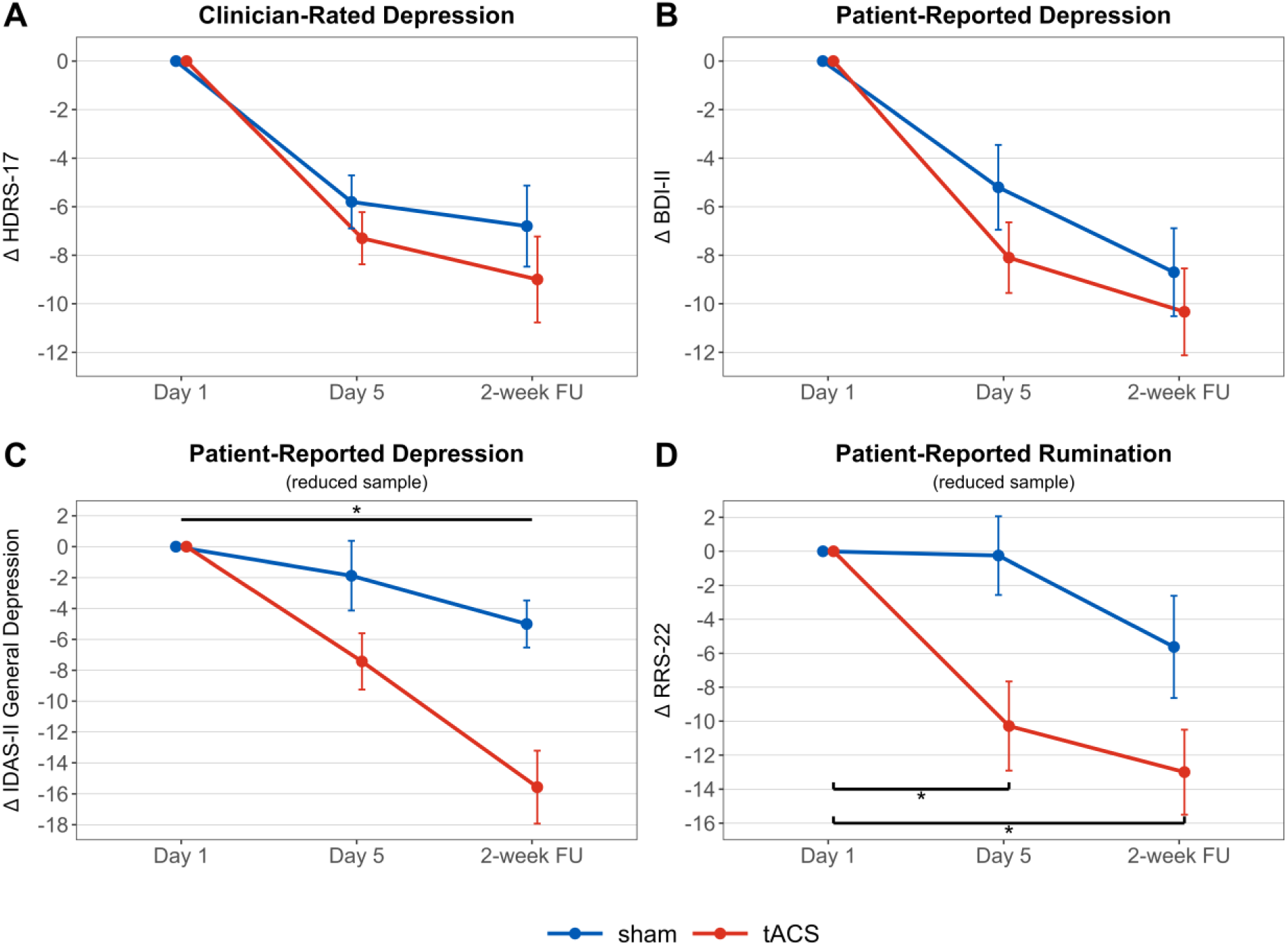
Clinical Outcomes. Difference scores are shown for Day 5 minus Day 1 (D5–D1) and 2-week follow-up minus Day 1 (FU–D1). Error bars indicate the standard error of the mean. **A:** Hamilton Depression Rating Scale (HDRS-17). **B:** Beck Depression Inventory (BDI-II). **C:** Inventory of Depression and Anxiety Symptoms (IDAS), General Depression subscale, shown for the reduced sample; * indicates a significant condition × time interaction (p = 0.002). **D:** Ruminative Responses Scale (RRS-22), shown for the reduced sample; * indicates significant within-group reductions in rumination in the tACS group from D1 to D5 (p = 0.034) and from D1 to FU (p = 0.001), in the absence of a significant condition × time interaction.

In the reduced sample, linear mixed-effects models revealed a significant condition × time interaction for IDAS General Depression scores, F(2, 31.86) = 6.01, p = .006, ηp² = .27, alongside a main effect of time, F(2, 31.86) = 24.56, p < .001, ηp² = .61. This interaction was driven by a 10.4-point greater reduction from Day 1 to 2-week follow-up in the tACS group relative to sham, reflecting an adjusted difference in change estimated from the mixed-effects model (β = −10.40, SE = 3.00, p = .002). For the RSS, there was a significant main effect of time, F(2, 27.86) = 11.28, p < .001, ηp² = .45, but no condition × time interaction, F(2, 27.86) = 2.13, p = .137, ηp² = .13. Post hoc comparisons based on model-estimated marginal means (Holm-corrected) indicated a significant reduction from Day 1 to 2-week follow-up in the tACS group (Δ = −11.85, SE = 3.07, p = .001), whereas the reduction in the sham group (Δ = −6.61, SE = 2.88) did not reach significance (p = .083) (Figure 5D). Results and figures for the quality of life and anhedonia questionnaires can be found in the supplement (Figure S4).

### Blinding Assessment

Participant blinding was assessed after the final stimulation session using a single dichotomous question (yes/no) asking whether participants believed they had received active tACS. One sham participant’s response was lost due to a data storage failure. In the tACS group, 6 of 10 (60%) participants believed they had received active tACS, while 6 of 9 (66.67%) in the sham group reported believing they had received active tACS. Fisher’s exact test showed no significant association between participants’ guesses and their actual condition (p = 1), indicating successful blinding.

### Tolerability

No serious adverse events occurred. Adverse events, collected semi-structurally at Day 5 and follow-up, largely reflected expected stimulation-related sensations; 137 of 166 events (82.5%) were rated as mild and 29 (17.5%) moderate in severity, with none rated as severe. The most frequently reported stimulation-related sensations during the intervention week were tingling (mean occurrences per participant over 5 days: 4.0 in tACS vs 3.2 in sham), sleepiness (2.6 vs. 2.9), and burning sensation (1.7 vs. 2.4), with no between-group differences in frequency (all p > .05; Table S2/Figure S5). Average severity ratings were low and did not differ between conditions (tACS: M = 0.36, SD = 0.64; sham: M = 0.35, SD = 0.58; Table S3/Figure S6).

## Discussion

We performed a randomized controlled trial to investigate the network neurophysiology of tACS in MDD to examine the mechanism of action for the reduction in depression symptoms. Five-day 10 Hz tACS was associated with frequency-specific changes in alpha power, characterized by a delayed reduction in posterior 10 Hz power and a relationship between IAF power modulation and clinical improvement in the tACS group.

### Suppression of Posterior 10 Hz Alpha Oscillations

High-density EEG revealed suppression of 10 Hz power over posterior electrodes at two-week follow-up. This effect was frequency-specific in that it was confined to the stimulation frequency, but its delayed emergence and posterior distribution indicate that it does not simply reflect local entrainment at the stimulation sites. One possible explanation is that interindividual variability in IAF may have limited frequency matching during stimulation [10,12], potentially contributing to the absence of significant effects at Day 5. Consistent with this possibility, natural frequencies reported for posterior alpha generators have been estimated to cluster around 10 Hz in occipital and precuneus regions [28], which may favor engagement of posterior rhythms by a 10 Hz tACS. In addition, posterior electrodes, where alpha power is typically highest, offer a superior signal-to-noise ratio, potentially making stimulation-related changes more detectable there than over prefrontal sites.

The absence of significant effects during the intervention week, alongside effects emerging at follow-up, suggests that repeated stimulation may induce delayed changes in oscillatory activity. Given that EEG was recorded at rest prior to stimulation, the current analysis is not sensitive to immediate entrainment effects, and the observed modulation likely reflects processes evolving across repeated sessions. Such temporal profiles are compatible with plasticity-related mechanisms, including spike-timing–dependent processes [13,29,30]. Notably, despite the bifrontal–central stimulation montage, effects were most evident over posterior electrodes, compatible with potential top-down control of prefrontal regions on occipital–parietal alpha oscillations ( 31,32).

Posterior alpha oscillations are associated with sensory gating, attentional allocation, and internally oriented processing [4,6], and alterations have been reported in depression [7], where they have been linked to excessive internally directed cognition [33]. At the network level, converging EEG–fMRI evidence relates alpha dynamics to intrinsic network organization, including the default mode network (DMN), although the direction of these relationships can vary across network components [34–36]. Resting-state fMRI studies in MDD further report increased within-DMN connectivity and altered coupling between the DMN and fronto-parietal control network (FPN) [37], consistent with reduced top-down regulatory control over internally generated cognitive processes [38]. Notably, recent tACS–fMRI studies demonstrate that modulation of alpha oscillations can alter DMN connectivity, providing causal support for a link between alpha activity and intrinsic network dynamics [39,40]. Within this framework, the observed posterior alpha modulation may reflect a shift in the balance between internally oriented and control-related processing, although this interpretation remains indirect.

### IAF Power Suppression Predicts Depression Improvement

Both ROI and topographical analyses demonstrated that reductions in IAF power were associated with improvement in depressive symptoms, an effect observed in the tACS group only. Similar associations have been reported across multiple treatment modalities, including tACS [14,41], antidepressants [42], psychotherapy [43], and esketamine [44]. The present findings extend this literature by demonstrating a dissociation between stimulation-related and symptom-associated effects on alpha oscillations. While alpha suppression was observed at stimulation frequency, its association with clinical improvement was specific to modulation at the IAF. This relationship was temporally delayed and spatially distributed both in prefrontal regions targeted directly by stimulation and across broader cortical areas, supporting the role of IAF power suppression as a candidate marker of target engagement.

Specifically, depression symptom improvement was linked to changes in power at the endogenous oscillatory peak, the IAF, rather than at the 10 Hz stimulation frequency. In dynamical systems, the natural resonance frequency defines the frequency at which a system is most responsive to external input [45]. The IAF reflects the dominant alpha rhythm and may approximate this intrinsic resonance frequency [46]. Accordingly, while stimulation at 10 Hz may induce measurable effects at the stimulation frequency, clinically relevant effects may depend on the degree to which stimulation engages the dominant intrinsic alpha rhythm. Consistent with this interpretation, prior studies demonstrate enhanced antidepressant efficacy when rTMS is applied near the IAF [47,48], supporting frequency-individualized stimulation approaches [41,49].

The relationship between IAF power modulation and clinical improvement was both temporally and spatially structured. Early suppression during the intervention week predicted later symptom improvement from a centro-parietal cluster, whereas longer-term FU-D1 reductions were associated with a more anterior distribution encompassing frontal and left parietal electrodes. These dissociable patterns suggest a posterior-to-anterior shift in alpha modulation over time, with early centro-parietal and later frontal–parietal changes reflecting distinct components of treatment-related neural dynamics.

Given the spatial constraints of scalp EEG and the influence of volume conduction, anatomical inferences remain indirect. The early central–parietal distribution is consistent with dominant posterior alpha generators, whereas the later frontal–parietal pattern aligns with regions implicated in top-down regulatory processes [50]. This dissociation is consistent with evidence that posterior alpha oscillations preferentially index internally directed and self-referential processing, whereas frontal alpha dynamics are more closely linked to cognitive control and top-down regulation [51,52].

At the systems level, this posterior-anterior alpha shift aligns with fMRI models of depression implicating dysregulated interactions between the DMN and the FPN [37,53,54]. Early posterior alpha suppression may reflect modulation of DMN-dominant intrinsic activity, potentially reducing the dominance of internally oriented processing. Subsequent frontal–parietal alpha changes may index progressive recruitment of FPN-mediated control processes. While these interpretations remain indirect, they suggest that clinically relevant effects may depend on how stimulation reshapes the balance between intrinsic and control-related network dynamics. However, testing whether these sensor-level alpha changes map onto DMN–FPN connectivity will require concurrent or longitudinal fMRI.

These findings suggest that aligning stimulation frequency with the IAF is critical for effective target engagement [41,55]. Future studies should determine whether this approach can be further refined by considering region-specific alpha peaks, rather than relying on a global IAF estimate that is largely driven by posterior activity [28]. This may be particularly relevant for individuals with lower alpha frequencies, which can be expressed more anteriorly and do not necessarily conform to the conventional 8–12 Hz alpha band definition. In parallel, optimization of stimulation protocols may also benefit from adjustments in dosage and temporal precision. State-dependent or closed-loop approaches may further enhance target engagement by adapting stimulation to ongoing brain states [49,56].

### Clinical Results

Numerically, 10 Hz tACS led to a 2.2-point greater reduction on the HDRS-17 compared with sham and was accompanied by a significant condition × session interaction on the IDAS-II General Depression subscale in a reduced sample. Similar magnitudes of HDRS-17 reduction have been reported in trials of recently FDA-cleared at-home brain stimulation devices [57,58], albeit following multi-week treatment protocols. Importantly, substantial symptom improvement was also observed in the sham group, consistent with non-specific treatment effects. However, the association between alpha modulation and clinical improvement was specific to the active condition, suggesting that electrophysiological changes do not merely reflect general symptom improvement but index stimulation-specific target engagement. Together, the convergence of clinical and mechanistic findings supports further evaluation in larger, adequately powered randomized studies.

### Limitations

The modest sample size limits statistical power, particularly for detecting clinical effects, and increases the risk of inflated effect sizes in correlation analyses. Although the analyses were guided by mechanistic hypotheses, the findings should be interpreted with caution. The two-week follow-up precludes inference about long-term durability. Stimulation was delivered at a fixed 10 Hz frequency using a bifrontal montage, limiting generalizability to frequency-individualized protocols or alternative electrode configurations. EEG analyses were conducted in sensor space, and anatomical inferences therefore remain indirect due to volume conduction. To address multiple comparisons in topographical analyses, statistical inference relied on cluster-based permutation testing. Lastly, baseline differences in episode duration and psychotherapy exposure may have influenced clinical outcomes.

## Conclusion

This study provides evidence that tACS can engage frequency-specific modulation of endogenous alpha activity as a mechanistically relevant pathway linking stimulation to clinical improvement in depression. By demonstrating that changes of IAF power predict symptom reduction, the findings help bridge the gap between target engagement and clinical outcome in non-invasive brain stimulation. The temporal and spatial structure of these effects further suggest a model of progressive network-level reorganization. Together, these results advance a framework for mechanistically informed and individualized tACS, with implications for optimizing stimulation parameters and guiding future efficacy trials.

## Data Availability

De-identified data produced in the present study are available from the authors upon reasonable request.

## Acknowledgements

Tobias Schwippel utilized ChatGPT to assist with language refinement.

## Author Contributions

**Tobias Schwippel:** Methodology, Data Curation, Investigation, Formal Analysis, Visualization, Writing – Original Draft. **Francesca Pupillo:** Data Curation, Investigation, Visualization, Writing – Review & Editing. **Hadden LaGarde:** Data Curation, Investigation, Visualization, Writing – Review & Editing. **Athena Stein:** Writing – Review & Editing. **Mengsen Zhang:** Supervision, Writing – Review & Editing **David Rubinow:** Supervision, Writing – Review & Editing. **Flavio Frohlich:** Conceptualization, Funding Acquisition, Resources, Supervision, Writing – Review & Editing.

All authors: Approved the final version of the manuscript.

