## Supplementary Material for "Suppression of Endogenous Alpha Power Predicts Clinical Response to 10 Hz tACS in Major Depressive Disorder: A Double-Blind Randomized Controlled Trial"

### Supplementarty Methods

#### CONSORT 2025 Flow Diagram

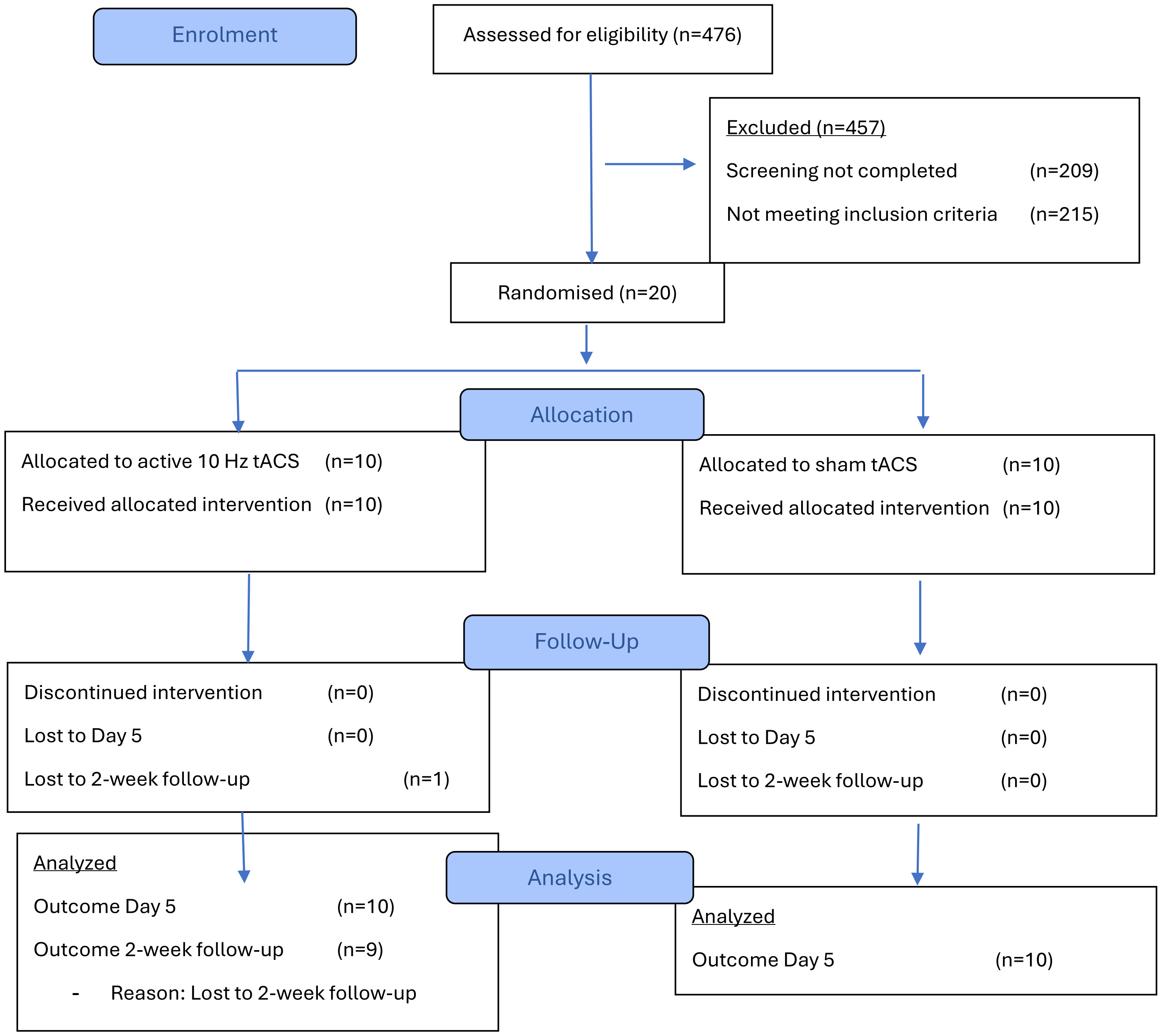

Hopewell, S., Chan, A. W., Collins, G. S., Hróbjartsson, A., Moher, D., Schulz, K. F., ... & Boutron, I. (2025). CONSORT 2025 statement: updated guideline for reporting randomised trials. The Lancet, 405(10489), 1633-1640. [1]

**Figure S1. Consort Flow Chart**

#### Statistical Analyses

All analyses were performed in R 4.2 [2] and MATLAB R2025a (The MathWorks, Inc., MA, USA). Statistical significance was set at $\alpha = 0.05$ (two-tailed). EEG analyses focused on PSD at 10 Hz and IAF. Due to the small sample size, nonparametric tests were used for group comparisons of EEG data and correlations.

Primary Analyses. Prefrontal alpha PSD difference scores within the intervention week (D5-D1) and from D1 to FU were compared between tACS and sham using Wilcoxon rank-sum tests. Spearman rank correlations assessed the relationship between PSD and HDRS-17 difference scores at the same timepoints. These ROI-based analyses correspond to the pre-specified primary outcomes registered on ClinicalTrials.gov (NCT03994081).

Secondary Analyses. Beyond the pre-registered primary outcomes, exploratory analyses included: (1) whole-head topographical PSD changes from D1 to D5 and from D1 to FU, (2) correlations between these PSD changes and HDRS-17 changes, including associations between PSD changes during the intervention week and HDRS-17 changes to FU, and (3) clinical outcome measures across all timepoints. The detailed statistical analysis plan for these additional outcomes is presented below.

(1) Topographical Analyses: Alpha PSD Change. To complement the ROI-based results, whole-head analyses across all 91 channels of alpha PSD and HDRS-17 changes were conducted. First, for each participant and condition, PSD difference scores at each electrode were calculated between timepoints. Statistical comparisons of PSD change between tACS and sham groups were performed at each electrode using nonparametric cluster-based permutation testing (MASSUNI toolbox, clust_perm2 function)[3]. Electrodes exceeding an initial uncorrected threshold of $p < 0.05$ were eligible for cluster formation. Significance was determined using 10,000 permutations with a fixed random seed, and family-wise error rate (FWER) correction was applied across the scalp. Cluster mass and FWER-corrected p-values are reported for each significant cluster.

(2) Topographical Analyses: Alpha PSD and HDRS-17 Change. Channel-wise correlations between PSD and HDRS difference scores were computed separately for tACS and sham using Spearman correlations. Multiple comparisons across electrodes were controlled using the false discovery rate (FDR).

(3) Clinical Outcomes. HDRS-17 and BDI II were analyzed using two-way repeated measures ANOVA (RM-ANOVA) with stimulation condition (tACS vs. sham) as the between-subject factor and time (D1, D5, FU) as the within-subject factor. When the assumption of sphericity was violated, Greenhouse–Geisser corrections were applied. Post hoc comparisons were

conducted using Tukey's HSD tests with adjustment for multiple comparisons. Partial eta-squared ($\eta^2_p$) values were reported as effect size. Treatment response was defined as a ≥50% reduction in HDRS-17 total score from baseline, and remission was defined as an HDRS-17 total score < 8 at 2-week FU. Between-group differences in response and remission rates were analyzed using two-sided Fisher's exact tests.

Clinical outcomes with missing observations were analyzed using linear mixed-effects models with condition, time (modeled as a categorical factor), and their interaction as fixed effects, and participant-level random intercepts, allowing inclusion of all available data. Models were estimated using maximum likelihood, and p-values were obtained using Satterthwaite's approximation [4]. Where applicable, post hoc contrasts were computed from model-estimated marginal means and adjusted for multiple comparisons using the Holm method [5].

(4) Stimulation-related Sensations. Anticipated stimulation-related sensations (tingling, warming, scalp pain, headache, neck pain, ringing, trouble concentrating, dizziness, palpitations, nausea, flickering lights) were assessed after each intervention session and rated on a 4-point Likert scale (0–3; absent to severe). Between-group differences in occurrence frequencies and mean overall severity were analyzed using independent-samples tests (Fisher's exact tests for occurrence; independent t tests for mean severity).

### Supplementarty Results

#### Demographic Information

Table S1. Demographics

| | tACS | sham |
|---|---|---|
| Age (M, SD) | 39.60 (20.55) | 37.60 (16.55) |
| Gender (n) | | |
| Female | 8 | 8 |
| Male | 2 | 2 |
| Race (n) | | |
| White | 8 | 7 |
| Asian | 0 | 1 |
| Black or African American | 1 | 1 |
| American Indian/Alaskan Native | 1 | 0 |
| Mixed Race | 0 | 1 |
| Ethnicity (n) | | |
| Hispanic or Latino | 0 | 1 |
| Not Hispanic or Latino | 10 | 9 |
| Current Episode Duration (months) | 3.15 (1.56) | 10.78 (8.44) |
| Medication (M, SD) | | |
| Antidepressants | 1.30 (0.67) | 1.20 (1.03) |
| Stimulants | 0.00 (0.00) | 0.30 (0.48) |
| Mood Stabilizers | 0.00 (0.00) | 0.20 (0.42) |
| Buspirone | 0.10 (0.32) | 0.10 (0.32 |
| Hydroxyzine | 0.20 (0.42) | 0.00 (0.00) |
| Antipsychotics | 0.20 (0.42) | 0.20 (0.42) |
| Current Psychotherapy (n) | | |
| Yes | 2 | 8 |
| No | 8 | 1 |
| Maudsley (M, SD) | 5.60 (0.84) | 6.00 (1.15) |
| HDRS-17 (M, SD) | 18.50 (3.81) | 17.10 (5.49) |

**Table S1. Demographics.** Maudsley: Maudsley Staging Method; HDRS-17: Hamilton Depression Rating Scale 17-item; M: mean; SD: standard deviation; n: count

#### EEG Target Engagement (IAF PSD)

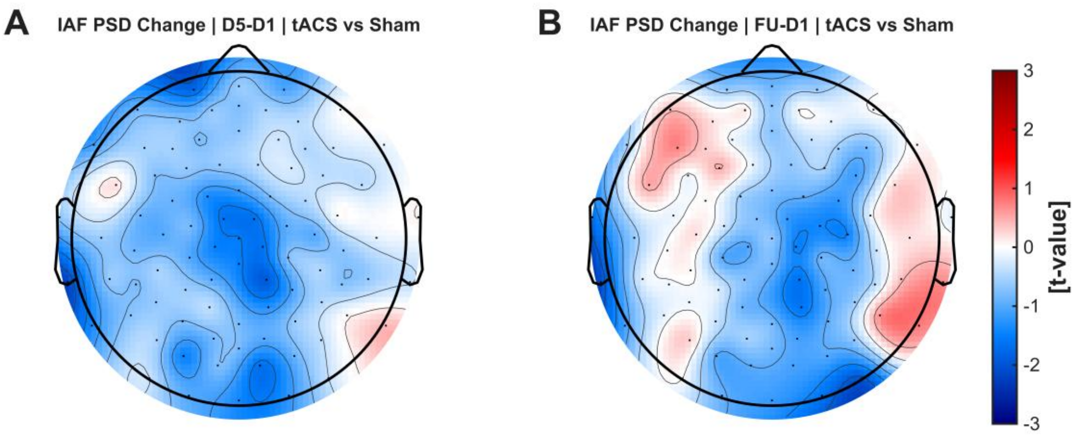

**Figure S2. Target Engagement: Topographical Analysis of IAF Hz PSD Modulation.**
Scalp distributions of t-values derived from nonparametric cluster-based permutation tests comparing PSD change between tACS and sham (double contrast: ΔtACS − Δsham). **A:** Change of IAF PSD from D1to D5 (intervention week). **B:** Change of IAF PSD from D1 to two-week FU. Colored maps display channel-wise t-statistics, scaled symmetrically around zero. Electrodes marked with white circles denote channels belonging to clusters surviving cluster-level FWER correction ($p < .05$). Negative t-values indicate greater reductions in IAF PSD in the tACS group relative to sham. Black dots represent EEG electrode locations. Abbreviations: IAF: individual alpha frequency; PSD: power spectral density; tACS: transcranial alternating current stimulation; D1: Day 1; D5: Day 5; FU: 2-week follow-up

#### EEG/HDRS Target Validation (10 Hz PSD)

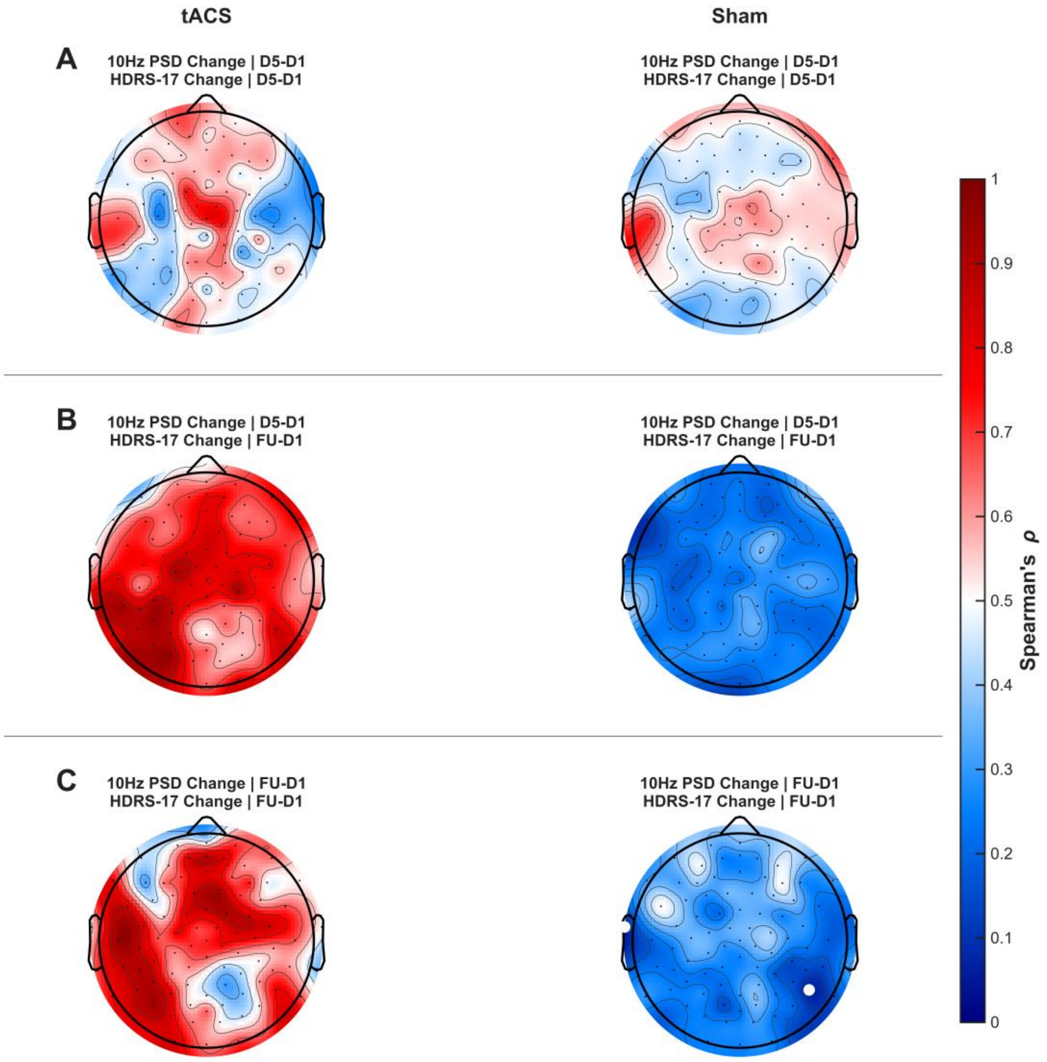

**Figure S3. Target Validation: Early and Sustained 10 Hz PSD Reduction Correlates with Symptom Improvement.**
Topographical maps depict Spearman correlation coefficients between changes in 10Hz PSD and changes in HDRS-17 across EEG channels, shown separately for tACS and sham groups. Colors indicate correlation coefficients, with positive values reflecting greater symptom reduction associated with larger PSD decreases. White circles indicate FDR-corrected significant channels (q < .05). Black dots indicate EEG electrode locations. **A:** Correlation between 10 Hz PSD change during the intervention week (D5–D1) and concurrent HDRS-17 change (D5–D1). **B:** Correlation between 10 Hz PSD change during the intervention week (D5–D1) and subsequent clinical improvement at 2-week follow-up (FU–D1). **C:** Correlation between sustained 10 Hz change from D1 to FU and HDRS-17 change from D1 to FU.
<u>Abbreviations</u>: PSD: power spectral density; tACS: transcranial alternating current stimulation; D1: Day 1; D5: Day 5; FU: 2-week follow-up

#### Secondary Clinical Outcomes

For secondary clinical outcomes (Q-LES-Q-SF and SHAPS), linear mixed-effects models with fixed effects of condition (sham vs. tACS), time (D1, D5, FU), and their interaction, and a participant-level random intercept were applied. Models were fitted to the raw observed scores rather than baseline-corrected difference values.

Quality of Life (Q-LES-Q-SF). For Q-LES-Q-SF, the sham group included 10 participants at Day 1, 8 at Day 5, and 8 at follow-up, while the tACS group included 10 participants at Day 1, 7 at Day 5, and 7 at follow-up, resulting in 15–20 participants contributing per time point.

The model revealed a significant main effect of time, $F(2, 33.21) = 10.33$, $p < .001$, $\eta p^2 = .38$, indicating improvement in quality of life across assessments. There was no main effect of condition, $F(1, 21.10) = 1.54$, $p = .228$, $\eta p^2 = .07$, and no condition × time interaction, $F(2, 33.21) = 0.08$, $p = .923$, $\eta p^2 = .005$.

Model-estimated marginal means indicated that Q-LES-Q-SF scores increased over time in both groups. In the sham group, scores increased from 42.3% at D1 to 53.2% at D5 (+10.9 percentage points) and 49.9% at follow-up (+7.5 percentage points). In the tACS group, scores increased from 38.6% at D1 to 48.2% at D5 (+9.6 percentage points) and 44.3% at follow-up (+5.8 percentage points). Post hoc comparisons based on model-estimated marginal means (Holm-adjusted) showed a significant increase from D1 to D5 in both sham (p = .008) and tACS (p = .034). The D1 to follow-up comparison showed a trend in the sham group (p = .066) but was not significant in the tACS group (p = .235). No between-group differences were observed at any time point (all p > .29).

Anhedonia (SHAPS). For SHAPS, the sham group comprised 10 participants at Day 1 and 8 participants at both Day 5 and follow-up. In the tACS group, 9 participants were available at Day 1 and 7 participants at both Day 5 and follow-up, resulting in 15–19 participants contributing per assessment.

The model revealed a significant main effect of time, $F(2, 31.72) = 5.92$, $p = .007$, $\eta p^2 = .27$, indicating a reduction in anhedonia across assessments. There was no main effect of condition, $F(1, 19.81) = 0.33$, $p = .572$, $\eta p^2 = .02$, and no condition × time interaction, $F(2, 31.72) = 0.72$, $p = .496$, $\eta p^2 = .04$.

Model-estimated marginal means indicated that SHAPS scores decreased from 6.20 at D1 to 4.21 at D5 (−1.99 points) and 4.59 at follow-up (−1.61 points) in the sham group. In the tACS group, scores decreased from 7.00 at D1 to 4.36 at D5 (−2.65 points) but increased to 6.36 at follow-up (−0.64 points relative to baseline). Post hoc comparisons based on model-estimated marginal means (Holm-adjusted) did not reveal statistically significant changes within either group. In the sham group, neither D1–D5 (p = .157) nor D1–FU (p = .225) contrasts were significant. In the tACS group, the D1–D5 contrast approached significance (p = .051), but all other comparisons were non-significant. No between-group differences were observed at any time point (all p ≥ .36).

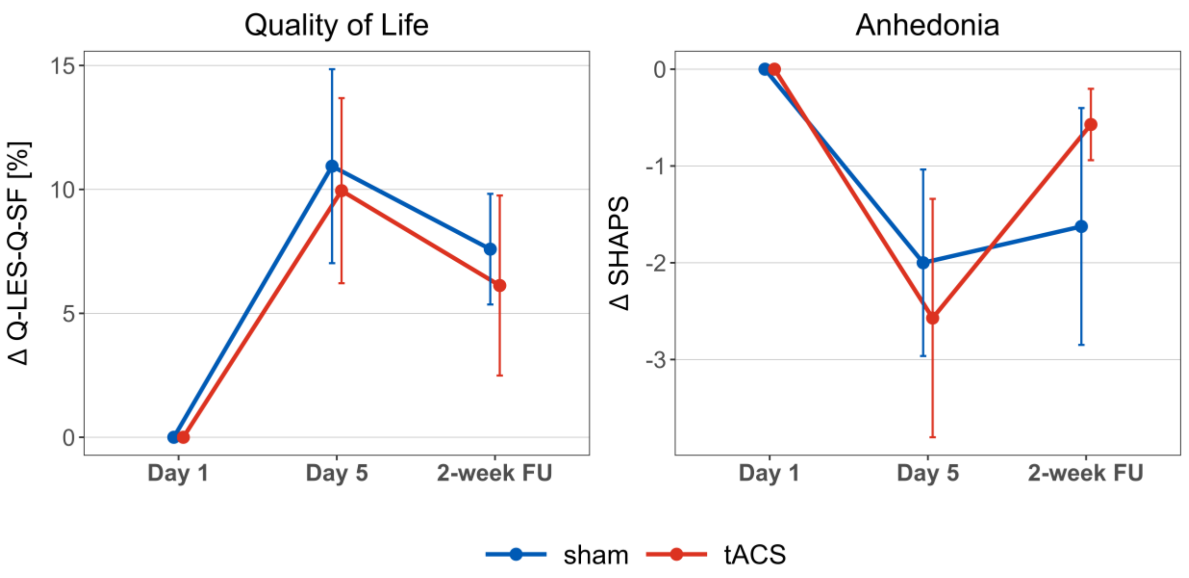

**Figure S4. Secondary Clinical Outcomes in the reduced sample.**
Changes in quality of life and anhedonia. Mean changes relative to Day 1 (D1) are shown for Day 5 (D5–D1) and the 2-week follow-up (FU–D1) in the tACS and sham groups. **A:** Quality of Life Enjoyment and Satisfaction Questionnaire-Short Form (Q-LES-Q-SF; percent score). **B:** Snaith–Hamilton Pleasure Scale (SHAPS). Points represent raw observed group means and error bars indicate the standard error of the mean.

**Stimulation-Related Sensations: Occurrence**

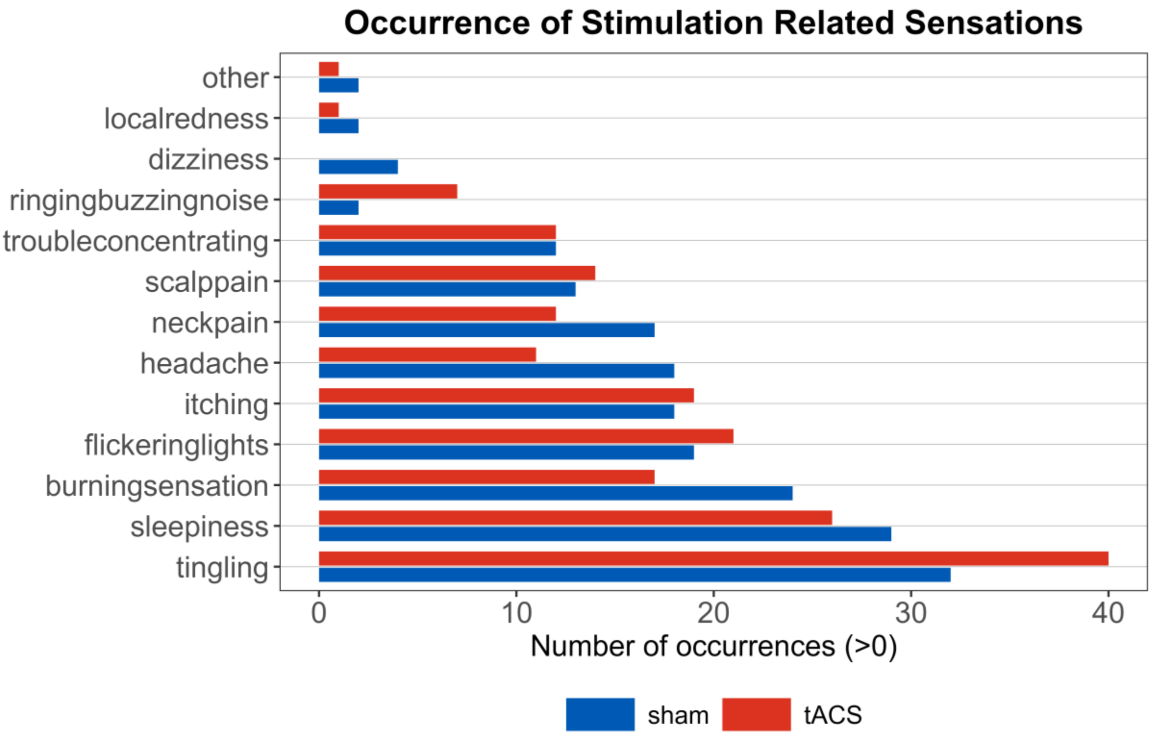

**Figure S5.** Occurrence of Stimulation-Related Sensations.
Counted for each participants on each of the five intervention days.

Table S2. Test Statistic for Occurrence of Stimulation-Related Sensations

| Symptom | M [sham] | SD [sham] | M [tACS] | SD [tACS] | t | df | p |
|---|---|---|---|---|---|---|---|
| tingling | 3.2 | 1.93 | 4 | 1.05 | -1.15 | 13.92 | 0.27 |
| sleepiness | 2.9 | 1.97 | 2.6 | 1.71 | 0.36 | 17.66 | 0.721 |
| burning sensation | 2.4 | 1.96 | 1.7 | 1.89 | 0.81 | 17.98 | 0.426 |
| flickering lights | 1.9 | 2.18 | 2.1 | 1.85 | -0.22 | 17.54 | 0.828 |
| itching | 1.8 | 1.81 | 1.9 | 1.91 | -0.12 | 17.95 | 0.906 |
| headache | 1.8 | 1.69 | 1.1 | 1.45 | 1 | 17.6 | 0.333 |
| neck pain | 1.7 | 1.95 | 1.2 | 1.4 | 0.66 | 16.34 | 0.519 |
| scalp pain | 1.3 | 1.57 | 1.4 | 1.78 | -0.13 | 17.72 | 0.895 |
| trouble concentrating | 1.2 | 1.62 | 1.2 | 1.48 | 0 | 17.85 | 1 |
| ringing buzzing noise | 0.2 | 0.42 | 0.7 | 1.57 | -0.97 | 10.3 | 0.352 |
| dizziness | 0.4 | 0.7 | 0 | 0 | 1.81 | 9 | 0.104 |
| local redness | 0.2 | 0.63 | 0.1 | 0.32 | 0.45 | 13.24 | 0.662 |
| other | 0.2 | 0.42 | 0.1 | 0.32 | 0.6 | 16.69 | 0.557 |

Occurrence of each sensation was collected on each intervention day (D1-D5) for each participant. For occurrence, the severity of the sensation was disregarded. M: Mean; SD: Standard Deviation; t: test statistic from the independent samples t-test; df: degrees of freedom

#### Stimulation-Related Sensations: Severity

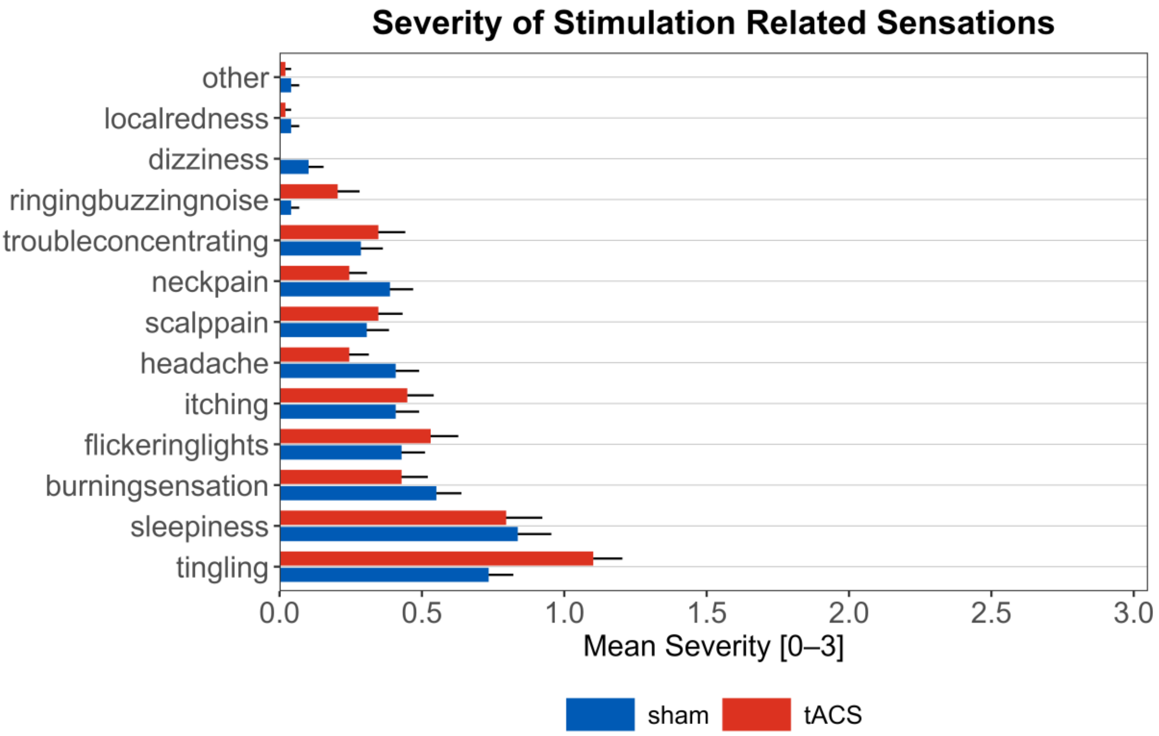

**Figure S6.** Severity of Stimulation-Related Sensations.
Rated on a 4-point Likert scale ranging from 0 (absent) to 3 (severe).

Table S3. Test Statistic for Severity of Stimulation Related Sensations

| Symptom | M [sham] | SD [sham] | M [tACS] | SD [tACS] | W | p |
|---|---|---|---|---|---|---|
| tingling | 0.74 | 0.48 | 1.1 | 0.54 | 32.5 | 0.197 |
| sleepiness | 0.84 | 0.64 | 0.8 | 0.65 | 51.5 | 0.939 |
| burning sensation | 0.56 | 0.49 | 0.42 | 0.51 | 58 | 0.564 |
| flickering lights | 0.42 | 0.5 | 0.54 | 0.52 | 43 | 0.611 |
| itching | 0.4 | 0.43 | 0.46 | 0.49 | 48.5 | 0.938 |
| headache | 0.41 | 0.38 | 0.24 | 0.32 | 64.5 | 0.272 |
| scalp pain | 0.3 | 0.4 | 0.34 | 0.43 | 47.5 | 0.874 |
| trouble concentrating | 0.28 | 0.37 | 0.35 | 0.6 | 47.5 | 0.875 |
| neck pain | 0.38 | 0.45 | 0.24 | 0.28 | 56.5 | 0.638 |
| ringing buzzing noise | 0.04 | 0.08 | 0.2 | 0.44 | 43 | 0.517 |
| dizziness | 0.1 | 0.19 | 0 | 0 | 65 | 0.078 |
| localredness | 0.04 | 0.13 | 0.02 | 0.06 | 50.5 | 1 |
| other | 0.04 | 0.08 | 0.02 | 0.06 | 55 | 0.583 |

Severity was rated on each intervention day (D1-D5) by each participant. Likert scale from 0 (absent) to 3 (severe). The statistics includes ratings of lowest severity (0). M: Mean; SD: Standard Deviation; W: Wilcoxon rank-sum test statistic.
